# Reversal of Epigenetic Age with Diet and Lifestyle in a Pilot Randomized Clinical Trial

**DOI:** 10.1101/2020.07.07.20148098

**Authors:** Kara N. Fitzgerald, Romilly Hodges, Douglas Hanes, Emily Stack, David Cheishvili, Moshe Szyf, Janine Henkel, Melissa W. Twedt, Despina Giannopoulou, Josette Herdell, Sally Logan, Ryan Bradley

## Abstract

Manipulations to set back biological age and extend lifespan in animal models are well established, and translation to humans has begun. The length of human life makes it impractical to evaluate results by plotting mortality curves, so surrogate markers of age have been suggested and, at present, the best established surrogates are DNA methylation clocks. Herein we report on a randomized, controlled clinical trial designed to be a first step in evaluating the effect of a diet and lifestyle intervention on biological age. Compared to participants in the control group (n=20), participants in the treatment group tested an average 3.23 years younger at the end of the eight-week program according to the Horvath DNAmAge clock (p=0.018). Those in the treatment group (n=18) tested an average 1.96 years younger at the end of the program compared to the same individuals at the beginning with a strong trend towards significance (p=0.066 for within group change). This is the first such trial to demonstrate a potential reversal of biological age. In this study, the intervention was confined to diet and lifestyle changes previously identified as safe to use. The prescribed program included multiple components with documented mechanistic activity on epigenetic pathways, including moderate exercise, breathing exercises for stress, and a diet rich in methyl donor nutrients and polyphenols.

## 1 INTRODUCTION

Advanced age is the largest risk factor for impaired mental and physical function and many non-communicable diseases including cancer, neurodegeneration, type 2 diabetes, and cardiovascular disease (Jin et al., 2015; Sen et al., 2016). The growing health-related economic and social challenges of our rapidly aging population are well recognized and affect individuals, their families, health systems and economies. Considering economics alone, using the Future Elderly Model, Goldman showed that delaying aging by 2.2 years (with associated extension of healthspan) could save $7 trillion over fifty years. This broad approach was identified to be a much better investment than disease-specific spending (Goldman, 2017). Thus, if interventions can be identified that extend healthspan even modestly, benefits for public health and healthcare economics will be substantial.

*DNA methylation* is the addition of a methyl group to cytosine residues at selective areas on a chromosome (e.g. CpG islands, shelf/shore, exons, open sea). Methylation constitutes the best-studied, and likely most resilient of many mechanisms controlling gene expression (Li & Zhang, 2014). Unique among epigenetic markers, DNA methylation can readily and cheaply be mapped from tissue samples. Of 20+ million methylation sites on the human genome, there are a few thousand at which methylation levels are tightly correlated with age. Currently, the best biochemical markers of an individual’s age are all based on patterns of methylation (Horvath & Raj, 2018). This has led some researchers to propose that aging itself has its basis in epigenetic changes (including methylation changes) over time (Field et al., 2018; Johnson et al., 2012; Mitteldorf, 2013; Rando & Chang, 2012).

As of this writing, the best-studied methylation-based clock is the multi-tissue DNAmAge clock (Horvath, 2013). At the time this study design was approved, there were few viable alternatives. Horvath’s DNAmAge clock predicts all-cause mortality and multiple morbidities better than chronological age. Methylation clocks (including DNAmAge) are based on systematic methylation changes with age, with about 60% of CpG sites losing methylation with age and 40% gaining methylation. This is distinct from stochastic changes, “methylation drift”, unpredictable changes which vary among individuals and cell-by-cell within individuals. Systematic methylation changes include hypermethylation in promotor regions of tumor suppressor genes (inhibiting expression) and hypomethylation promoting inflammatory cytokines (promoting expression). Saliva can be considered a good source of high-quality DNA, containing both white blood cells and buccal cells, and is a suitable tissue type to be assessed for the DNAmAge clock (Horvath, 2013; Langie et al., 2017).

The dietary recommendations employed as part of the treatment protocol for this study were based largely on biochemistry and generalized measures of health, because few dietary associations with the DNAmAge clock have yet been established. A modest, but significant, reduction in DNAmAge in individuals consuming a non-specific lean meat, fish and plant-based diet (as measured by blood carotenoids) has been observed (Quach et al., 2017). It is possible that changes of a greater magnitude require a more targeted approach. The dietary intervention used here was also plant-centered, but including a high intake of nutrients that are substrates or cofactors in methylation biosynthetic pathways (e.g. containing folate, betaine), ten-eleven translocation demethylase cofactors and modulators (e.g. alpha ketoglutarate, vitamin C and vitamin A) (Hore, 2017) and polyphenolic modulators of DNA methyl transferases (DNMT) (e.g. curcumin, epigallocatechin gallate (EGCG), rosmarinic acid, quercetin, luteolin). It also included limited nutrient-dense animal proteins (e.g. liver, egg). The diet restricted carbohydrates and included mild intermittent fasting, both designed to lower glycemic cycling. The diet was supplemented daily with a fruit and vegetable powder, also rich in polyphenolic modulators of DNMT activity, and a probiotic providing 40 million CFU of *Lactobacillus plantarum* 299v. *L. plantarum* has been shown to be a folate producer in the presence of para aminobenzoic acid (PABA) (Sybesma et al., 2003); it also has been demonstrated to alter gene expression (Hariri et al., 2015).

Lifestyle guidance in this study included a minimum of 30 minutes of exercise per day, at least 5 days per week at an intensity of 60-80 percent of maximum perceived exertion. Exercise is well-known to be broadly beneficial for almost every aspect of health and has been shown to extend mean lifespan in animal models. Exploration of the effect of exercise on the methylome has recently begun. For example, regular *tai chi* practice was associated with slowing of age-related DNA methylation losses in 500 women (Ren et al., 2012). In another study of 647 women, a lifelong history of exercise was associated with a similar endpoint (White et al., 2013). These results were not reported in terms of the Horvath clock, because it had not yet been developed. One systematic review of human studies found that regular, daily physical activity was associated with lower blood levels of homocysteine, which when elevated, suggests an insufficiency of methylation capacity (e Silva & da Mota, 2014). Excessive exercise may accelerate methylation aging, but this danger has only been observed in elite, competitive athletes (Spólnicka et al., 2018).

Twice-daily breathing exercises that elicit the Relaxation Response were prescribed for stress reduction. It was recently demonstrated that 60 days of relaxation practice designed to elicit the Relaxation Response, 20 minutes twice per day, could significantly reduce DNAmAge as measured by the Zbieć-Piekarska clock in their group of healthy participants (though not in their ‘patient’ group) (Pavanello et al., 2019). *Almost* a quarter of the DNAmAge CpG sites (85/353) are located in glucocorticoid response elements, pointing to a likely relationship between stress and accelerated aging. Cumulative lifetime stress has been shown to be associated with accelerated aging of the methylome (Zannas et al., 2015). Zannas et al. also reported that dexamethasone, a glucocorticoid agonist, can advance the DNAmAge clock and induce associated transcriptional changes. Dexamethasone-regulated genes showed enriched association of aging-related diseases, including coronary artery disease, arteriosclerosis and leukemias. Other findings include that PTSD contributes to accelerated methylation age (Wolf et al., 2016); and that greater infant distress (lack of caregiver contact) is associated with an underdeveloped, younger epigenetic age (Moore et al., 2017).

This study aimed to optimize sleep, with a recommendation for at least seven hours nightly. Seven hours is generally considered to be healthy (Panel et al., 2015), but the limited data on accelerated aging only relates to extremes of sleep deprivation. A (presumably transient) effect of sleep deprivation on genome-wide methylation patterns in blood has been demonstrated (Nilsson et al., 2016). Acceleration of the DNAmAge clock has been associated with insomnia in a sample of 2078 women (Carroll et al., 2017). Carskadon et al (2019) found an association between poor quality / fewer hours of sleep with age acceleration in a small sample of 12 female college students.

This multimodal (“systems”) intervention is reflective of a clinically-used approach that combines individual interventions, each of which carry evidence of favorable influence on the DNA methylome and of which several authors of this study have clinical experience of health benefits. Such interventions likely produce synergistic effects and reduce the possibility of negative effects from one disease-promoting input canceling out the benefits of another health-promoting input. Dietary and lifestyle interventions, as used here, target upstream influences that are generally considered safe, even over the long term.

By design, an important endpoint of this study was to be Horvath’s DNAmAge clock, to see if it could be potentially slowed or reversed. This is to say we have tentatively accepted the hypothesis that the methylation pattern from which the DNAmAge clock is computed is a driver of aging (and the chronic diseases of aging), thus we expect that attempting to directly influence the DNA methylome using diet and lifestyle to set back DNAmAge will lead to a healthier, more “youthful” metabolism. To date, one small pilot study (Fahy et al., 2019) has been reported to have set back the DNAmAge clock over the course of 12 months by 1.5 (plus the one-year duration of the study) years in humans, using a combination of growth hormone, metformin, DHEA and two dietary supplements. Herein we report comparable initial results based on diet and lifestyle interventions employed for eight weeks (preceded by a one-week washout period).

## 2 RESULTS

### 2.1 Methylation clock setback

Compared to participants in the control group (n=20), participants in the treatment group scored an average 3.23 years younger at the end of the eight-week program according to the Horvath DNAmAge clock (p=0.018). Those in the treatment group (n=18) scored an average 1.96 years younger, at the end of the program compared to the same individuals at the beginning with a strong trend towards significance (p=0.066 for within group change). Control participants scored an average of 1.27 years older at the end of the study period, though this within-group increase was not statistically significant (p= 0.153).

Comparison of DNAmAge change between treatment and control groups is shown in Figure 1 whereas within group changes for the treatment group are shown in Figure 2.

**Figure 1.**
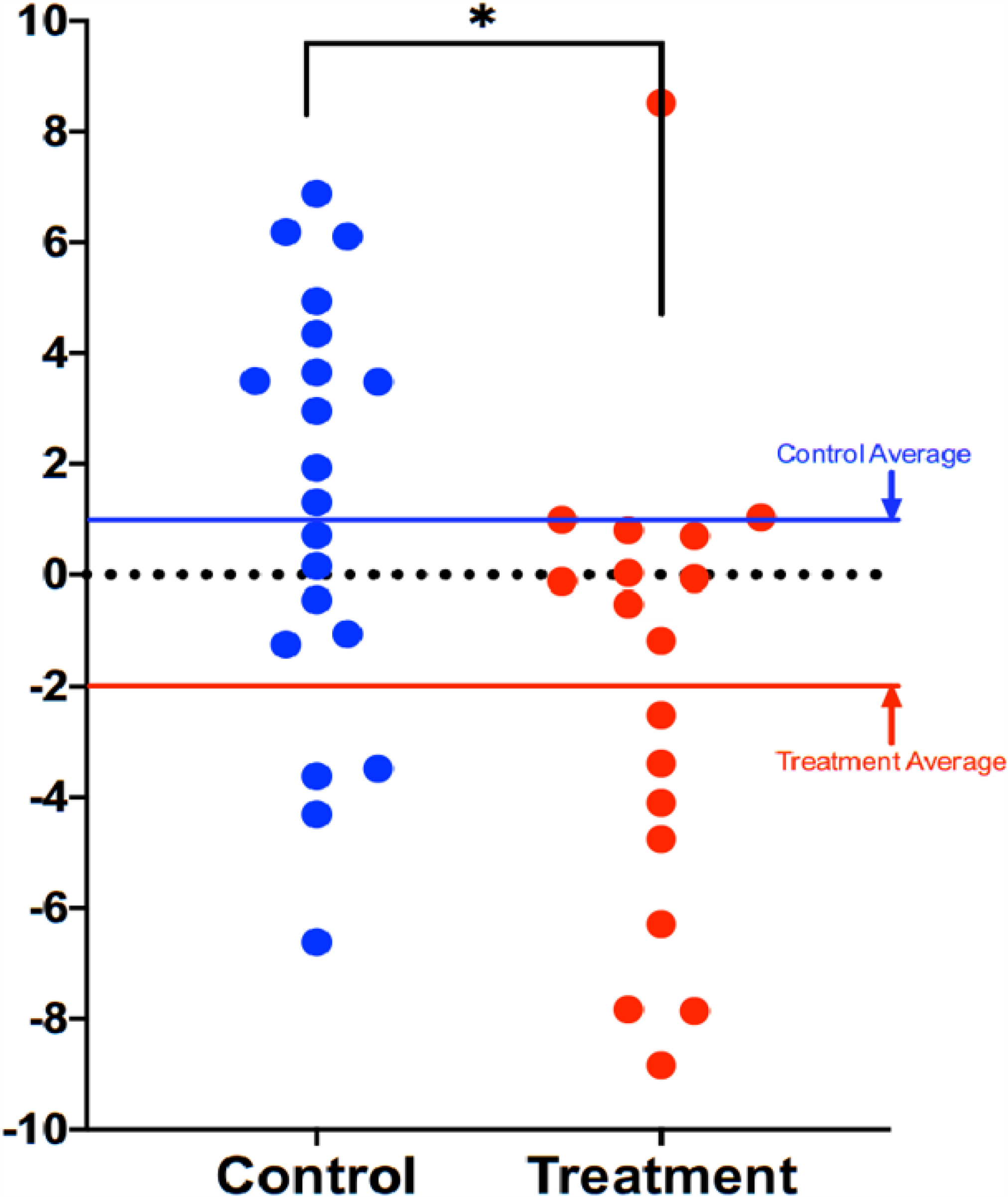
Comparison of DNAmAge change between treatment and control groups. Each dot is a subject, and the vertical axis represents difference in DNAmAge from the beginning to the end of the eight-week term. Participants scored an average 1.96 years younger, controls an average 1.27 years older. The age reduction of the treatment group strongly trended towards significance (p=0.066), while the age increase of the control group itself was not significant (p=0.153). The difference between control and treatment groups was significant at the level p=0.018 (unpaired two-tailed t-test). Long red and blue lines represent group averages (mean).

**Figure 2.**
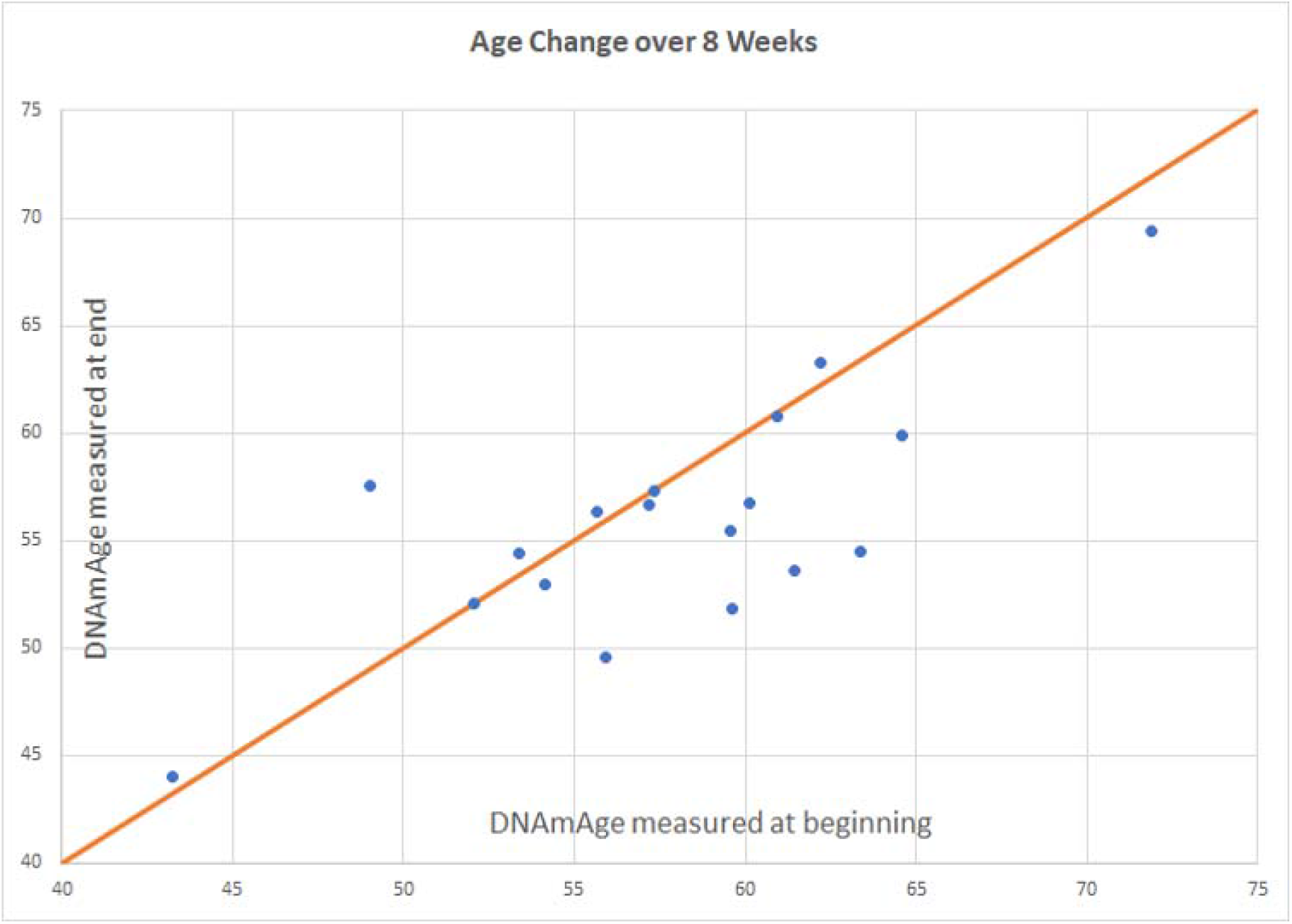
Intervention group age change. Participants scored an average of 1.96 years younger than baseline (p=0.066). Of 18 participants included in the final analysis, 8 scored age reduction, 9 were unchanged, and 1 increased in methylation age.

In both treatment and control groups, global average methylation stayed the same over the course of the study, with no net increase or decrease in the 353 sites that compose the Horvath clock.

## 3 DISCUSSION

### 3.1 Significance of Results

The significance of these findings is multi-factorial, but primarily as the first demonstration of potential reversal of epigenetic age in a randomized, controlled clinical trial, accounting for any normal variability in epigenetic methylation. Secondarily, this is the first report of a diet and lifestyle intervention reducing biologic aging. Notably, the scale of potential reduction, while modest in magnitude, may correlate with meaningful socioeconomic benefits, and appears to have the potential to be broadly achievable.

Published in the fall of 2019, the TRIIM study (Fahy et al., 2019) was the first demonstration of a set of interventions setting back the flagship Horvath clock DNAmAge (Horvath, 2013). In TRIIM, a one-year regimen of daily injection of growth hormone plus one prescription drug and three nutritional supplements was shown to set back the DNAmAge clock by 1.5 years in 9 middle-aged men (plus the 1-year study duration = 2.5 years). In the present study, age set-back was achieved in eight weeks, using less expensive, less invasive, and otherwise generally beneficial interventions known to have mechanistic plausibility for affecting methylation pathways.

### 3.2 Targeting Epigenetics with Diet

The seminal work of Waterland and Jirtle (Waterland & Jirtle, 2003) in the Agouti mouse model marked a defining point in our understanding that nutrition elements could so affect DNA methylation marks as to silence gene expression and dramatically alter phenotype. The power of nutrition to bring about transformative phenotypic changes has held up over the intervening years, most strongly in animal studies, but also in some limited human trials (Waterland & Jirtle, 2003). Both TRIIM and the present study were able to effect changes on the DNA methylome without extra-dietary supplementation of known methyl donor nutrients (e.g., folate, vitamin B12, choline, SAMe or betaine) Illustrating a far-reaching regulatory network on DNA methylation and representing a departure from previous studies that manipulated DNA methylation more directly with extra-dietary supplemental folate, B12 and other methyl donor nutrients (Pauwels et al., 2017; Sae-Lee et al., 2018; Waterland & Jirtle, 2003; Zhong et al., 2017).

### 3.3 Rationale for Not Using Supplemental Methyl Donor Nutrients

In designing the present study, extra-dietary supplementation of methyl donor nutrients was specifically avoided because a growing body of epidemiological evidence indicates potential long-term risks, to which the short-term studies were not sensitive. Although overall data are mixed, and certain conditions (e.g. pregnancy, macrocytic anemia, hyperhomocysteinemia, dietary limitations) often require extra-dietary supplementation, several trials have found a positive association between methyl donor supplementation and increased cancer risk: Published long-term follow up on 2,524 participants in the B-PROOF trial which assessed the effect of 2-3 years of daily supplementation with 400 mcg folic acid and 500 mcg vitamin B12 found an increased risk of overall cancer (HR 1.25, 95% CI 1.00-1.53), p=0.05) and colorectal cancer in particular (HR 1.77, 95% CI 1.08-2.90, p=0.02) (Oliai Araghi et al., 2019). A meta-analysis of 2 trials in Norway similarly reported that 800 mcg folic acid plus 400 mg vitamin B12 daily was associated with increased cancer outcomes and all-cause mortality (Ebbing et al., 2009). In contrast, dietary folate intake from food was found to be inversely associated with non-muscle-invasive bladder cancer progression in a study that also found higher recurrence for folic acid intake (Tu et al., 2018), and baseline dietary folate intake was inversely associated with prostate cancer risk in a trial that subsequently identified an increased risk of prostate cancer in the treatment arm that received 1 mg folic acid per day for 10 years (Figueiredo et al., 2009). Also relevant is the demonstration, albeit in a small study, adding dietary supplements of folic acid, vitamin B6 and vitamin B12 to a vitamin D plus calcium intervention increased biological aging (sex-adjusted odds ratio 5.26 vs vitamin D plus calcium alone) during a 1-year intervention (Obeid et al., 2018).

### 3.5 Polyphenols as Selective DNA Modulators

The DNAmAge clock is computed from some sites that increase and others that decrease methylation with age, so a net methylation increase would not necessarily be beneficial. Since this study targeted a healthy methylation pattern, not limited to increased methylation, the prescribed diet was rich in TET demethylase-associated nutrients (Hore, 2017) and specific plant polyphenols known to selectively regulate DNMT activity in addition to food-sourced methyl donors. It may be that these compounds assist in elevating methylation substrate and cofactor support from a risky ‘blunt instrument’ to ‘precision surgery’ on the DNA methylome by regulating where methyl groups are applied and removed.

### 3.6 Is Methylation Age Setback a Proxy for Reversed Senescence?

The DNA methylation⇒aging hypothesis has been well-articulated, though it not achieved universal acceptance. The idea that aging (like developmental biology) is rooted in epigenetics was first discussed around 2012 (Johnson et al., 2012; Mitteldorf, 2013; Rando & Chang, 2012), and has continued to gain adherents since (Barja, 2019; Horvath & Raj, 2018; Pal & Tyler, 2016). There may now be an emerging consensus that aging is driven by epigenetics (Sen et al., 2016) and that an epigenetic assault on aging is a feasible way to reduce multi-morbidities in an aging population (Jin et al., 2015). Even authors who have a historic commitment to a view of aging based in accumulated damage are acknowledging the practical potential of age reversal via epigenetics (Boyd-Kirkup et al., 2013; Hayano et al., 2019; Kane & Sinclair, 2019; West et al., 2019).

It is not yet fully established whether interventions that slow or reverse any of the “methylation clocks” necessarily curtail risks of age-related disease. This is a key question for a community of epidemiologists who are eager for reliable markers of aging that can measure the benefits of their interventions without waiting decades for morbidity and mortality to be measurably affected in experimental participants. For clinicians who follow the current research, they will certainly want to know how much confidence to lend to clinical studies in which the endpoint is methylation age.

### 3.5 Cautions and Future Directions

One significant limitation of this pilot trial is limited statistical power to due the relatively small sample size. Confirmation of these results is therefore needed in larger study groups and populations beyond middle-aged men.

The use of a multimodal intervention has advantages, as discussed above, however it also means that the contributions of any one component to the overall potential benefit of the intervention cannot be attributed to any individual element of the intervention. The combination of interventions used in this study may yet be improved upon and may be more impactful when further personalized. Future iterations of the intervention in continued clinical trials will attempt to optimize the program for efficacy, efficiency, scalability and affordability. An ever-evolving understanding of personalized application of such dietary and lifestyle interventions will likely lead to refinements to this kind of intervention that may further extend indicators of biological age.

Finally, it may be that emerging “omics” approaches continue to evolve our understanding of biological age prediction and reversal beyond DNA methylation alone (Lorusso et al., 2018). Integration of our future understanding of multi-omics data should therefore be considered in the future trials of candidate age-delaying interventions.

## 4. EXPERIMENTAL PROCEDURES

### 4.1 Study design and conduct

The trial design was approved by the Institutional Review Board of the National University of Natural Medicine (IRB number: RB100217) and registered at ClinicalTrials.gov (Identifier: NCT03472820). All trial procedures (visits, consent, randomization, and study visits) were conducted at the Helfgott Research Institute, 2220 SW 1^st^ Ave, Portland, OR 97201.

Participants were voluntarily recruited on a rolling basis from the general community surrounding Portland, OR, using flyers, online and newspaper advertising, and electronic newsletters. Volunteers completed telephone pre-screening followed by an initial onsite screening visit including confirmation of eligibility, willingness to follow the study protocol, and informed consent.

43 eligible adult males without history of recent or chronic disease, between the ages of 50-72, were recruited, consented, enrolled and randomized between March 2018 and August 2019. A CONSORT flow diagram is shown in Figure 3. Baseline characteristics are shown in Table 1. The age range of 50-72 was selected as a time when age-related vulnerabilities typically manifest, and the limitation to only male participants was to avoid the potential confounding factor of pre-, peri-, and post-menopausal sex hormone levels of the same age range in women.

**Table 1:** Baseline characteristics.

| Characteristics | Treatment Group<br>n = 21 |  | Control Group<br>n = 22 |  |
| --- | --- | --- | --- | --- |
|  | Value | % | Value | % |
| Age, years (mean ±SD) | 58.5 ± 6.12 | | 60.3 ± 6.68 | |
| **Race** |  |  |  |  |
| Black or African American | 0 | 0 | 2 | 9.1 |
| American Indian or Native Alaskan | 0 | 0 | 0 | 0 |
| Native Hawaiian or Other Pacific Islander | 0 | 0 | 0 | 0 |
| Asian or Asian American | 3 | 13.6 | 1 | 4.5 |
| White, Caucasian or European American | 18 | 81.8 | 18 | 81.8 |
| Caribbean Islander or African National | 0 | 0 | 0 | 0 |
| More than one race | 0 | 0 | 1 | 4.5 |
| Unknown | 0 | 0 | 0 | 0 |
| **Education Level** |  |  |  |  |
| Some high school | 0 | 0 | 0 | 0 |
| High school | 0 | 0.00 | 3 | 0.14 |
| Some university | 1 | 0.05 | 1 | 0.05 |
| 2 year university | 2 | 0.09 | 1 | 0.05 |
| 4 year university | 6 | 0.27 | 4 | 0.18 |
| Some graduate school | 2 | 0.09 | 4 | 0.18 |
| Graduate degree | 11 | 0.50 | 9 | 0.41 |

**Figure 3:**
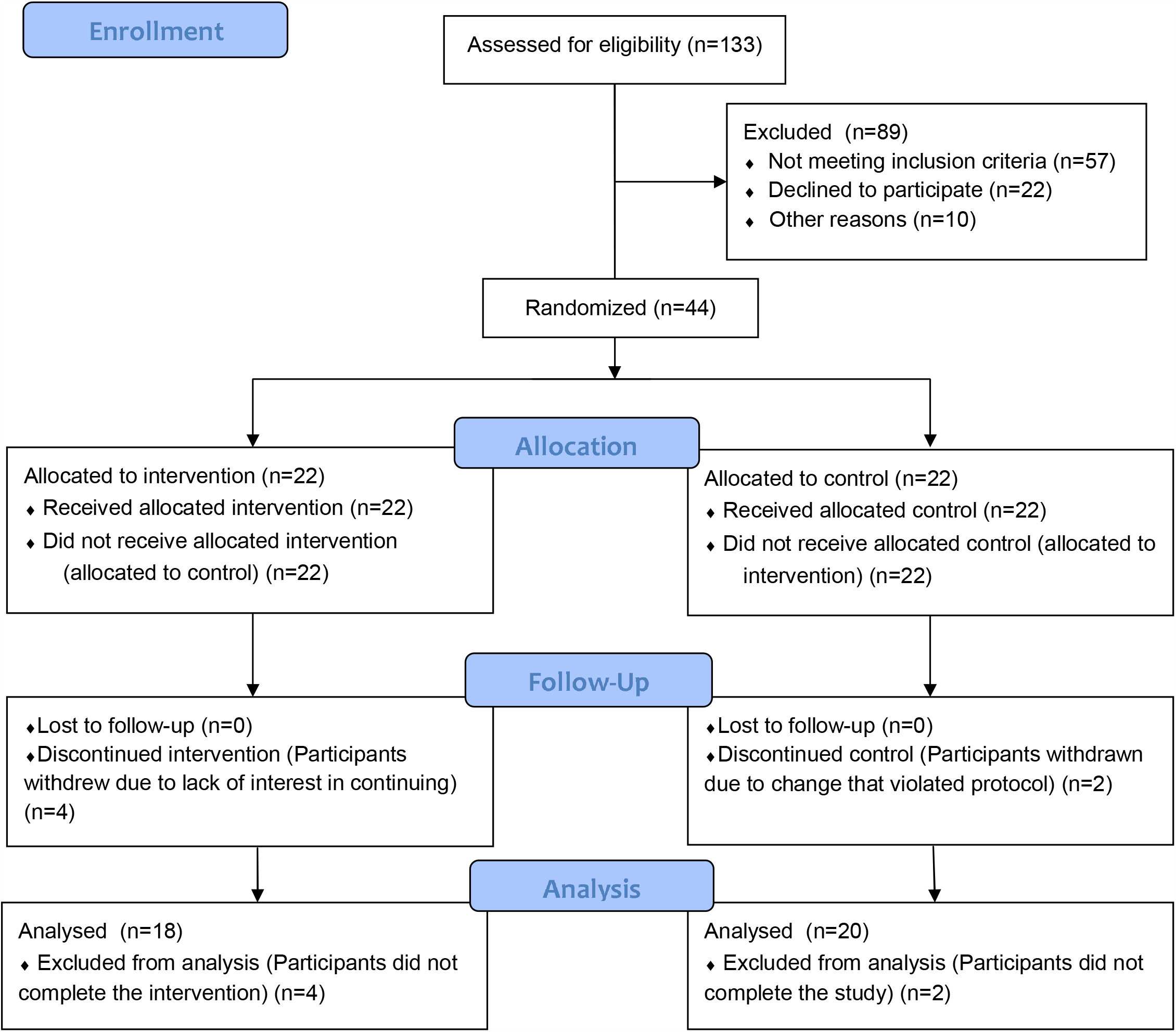
CONSORT 2010 Flow Diagram.

A 3-week washout period (with written instruction) was initiated for all participants involving discontinuation of any nutrition supplements or herbal products not prescribed by a licensed healthcare provider for a medical condition. Allowable exceptions for dietary supplementation included low dose supplements, such as a common “1-a-day” multivitamin/mineral (i.e. high potency, high dose multivitamin/mineral products were not allowable), and/or other supplements taken for prevention: fish oil (up to 1 gram/day), vitamin D (up to 6000 IU/day), vitamin C (up to 1g/day), vitamin E (up to 400 IU/day). Notably, these supplements were not recommended or prescribed, but participants already taking these products were allowed to continue them during the trial, thus any effects would be captured in their baseline assessments. Participants agreed to avoid/discontinue any recreational drugs/substances, as well as to consume no alcohol, nicotine, marijuana or cannabinoids at least 1 week before scheduled study visits.

Study participants were randomized at the baseline visit according to a randomization sequence (randomization.com). Allocation concealment was accomplished by opening sealed, signed envelopes prepared by research staff not associated with the trial which were only opened at randomization.

Initial instructions, including a recorded instructional webinar and electronic technology webinar were provided at visit 1. To allow time for participant education to occur, participants in the treatment group were instructed to begin the 8-week intervention protocol (including dietary, supplement, and lifestyle changes) starting one week after the baseline visit. Saliva samples were collected at each of the three study visits (baseline, week 5 and week 9).

An overview of the intervention is provided in Table 2. Two nutritional supplements (PhytoGanix® and UltraFlora® Intensive Care, Metagenics Inc., 25 Enterprise Aliso Viejo, CA 92656 USA) were distributed at visits 1 and 2. Unused doses were collected, counted and recorded at visits 2 and 3. Dosing adherence was verified by the retrospective review of returned doses, direct queries about each component of the intervention during study visits, and by frequent communication with trial participants.

**Table 2:** Summary of Dietary and Lifestyle Interventions*. ^A^Stress Management Recommendations were updated from the original Study Protocol as listed on ClinicalTrials.gov. All updates were IRB approved. *Patent pending.

| Intervention Category | Details |
| --- | --- |
| Dietary Prescription | *Guidance per week:*<br>**3 servings of liver**<br>• (1 serving = 3 oz)<br>• Preferably organic<br>**5-10 eggs**<br>• Ideally free-range, organic, omega-3 enriched<br>*Guidance per day:*<br>**2 cups of dark leafy greens**<br>• Measured raw, chopped, and packed<br>• Including kale, Swiss chard, collards, spinach, dandelion, mustard greens<br>• Does not include salad greens such as romaine, iceberg, Spring mix<br>**2 cups cruciferous vegetables**<br>• Measured raw, chopped, and packed<br>• Includes broccoli, cabbage, cauliflower, Brussels sprouts, bok choy, arugula, kale, mustard greens, watercress, rutabaga, kohlrabi, radish, Swiss chard, turnip<br>**3 additional cups colorful vegetables** of your choosing (excluding white potatoes, sweetcorn)<br>**1-2 medium beet**<br>**4 tbsp (1/4 cup) pumpkin seeds** (or pumpkin seed butter)<br>**4 tbsp (1/4 cup) sunflower seeds** (or sunflower seed butter)<br>**1+ serving methylation adaptogens, choose from:**<br>• 1/2 cup berries (wild preferred)<br>• 1/2 tsp rosemary<br>• 1/2 tsp turmeric<br>• 2 medium cloves garlic<br>• 2 cups green tea (brewed 10 minutes)<br>• 3 cups oolong tea (brewed 10 minutes)<br>**6 oz animal protein**<br>• Grass-fed, pastured, organic and hormone/antibiotic-free<br>**2 servings of low glycemic fruit**<br>*General guidance:*<br>**Organic** preferred over conventional<br>**Stay hydrated**<br>**Don't eat between** 7pm and 7am<br>**Include “healthy” oils**<br>• Balance types of fat<br>• E.g. coconut, olive, flaxseed and pumpkin seed oil<br>**Avoid** added sugar/candy, dairy, grains, legumes/beans<br>**Minimize** plastic food containers |
| Supplement Prescription | PhytoGanix®, a combination of organic vegetables, fruits, seeds, herbs, plant enzymes, prebiotics and probiotics at a dose of 2 servings daily, divided |
|  | UltraFlora® Intensive Care, containing *Lactobacillus plantarum* 299v at a dose of 2 capsules daily, divided |
| Exercise Prescription | Minimum of 30 minutes of exercise per day for at least 5 days per week, at an intensity of 60-80% of maximum perceived exertion |
| Sleep Prescription | Average a minimum of 7 hours of sleep per night |
| Stress Management Prescription <sup>A</sup> | Breathing exercise *Steps to Elicit the Relaxation Response* developed by Herbert Benson MD, twice daily |

Adherence to the program was supported by regular coaching sessions, delivered weekly during the first four weeks, and then at least every other week thereafter. Coaching sessions followed a pre-defined script that covered adherence to intervention guidelines and any changes to medications. A HIPAA-compliant electronic technology coaching tool was offered (MBody360, 640 Broadway 5A, New York, NY 10012) which contained reference instructions, meal planning ideas, optional recipes, and a shopping list.

Participants could also use this tool to communicate with their assigned coach in between scheduled coaching sessions. Email, web platform and/or phone communication were other options for participants unable or unwilling to use MBody360.

### 4.3 Determination of epigenetic age

#### Sample Handling

Saliva samples were stored at −70°C and remained frozen throughout the duration of the trial. Frozen samples were batch shipped overnight on dry ice to Yale University Center for Genome Analysis at the conclusion of clinical operations. Prior to shipment, sample IDs were assigned to plates such that each plate included a representative collection of samples from both allocation groups (treatment and control) and a distribution of samples from each trial visit, i.e., plates were not homogenous by group or by visit type, in an effort to equally distribute any random variability resulting from measurement across plates.

#### DNA Extraction

Oragene Saliva tubes were submitted and DNA extracted using the Perkin Elmer Chemagic 360 Instrument (kit# CMG-1081) following the manufacturer’s recommended protocol. An RNASE A digestion was added using 80uL of 4mg/uL Amercian Bioanalytical RNASE A (Part#AB12023-00100) after the 50 degree Celsius Oragene Saliva Incubation, before loading samples onto instrument.

#### Genomic DNA and RNA

The quality of the RNA/DNA was evaluated by: A260/A280 and A260/A230 ratios (as supplied by the NanoDrop 1000 Spectrophotometer), both of which should be > 1.8. The gel electrophoresis pattern was consistent with non-degraded samples.

#### Methods

Sample DNA was normalized to the recommended starting concentration, 1ug, for the Zymo EZ-96 DNA Methylation Kit (Cat No D5004). The samples underwent an overnight bisulfate conversion and were purified using the Zymo Methylation protocol. Samples proceeded directly to the Illumina Infinium HD assay (Illumina Methylation Epic Array Cat. No. WG-317-1001) to overnight whole genome amplification at 37 degrees Celsius. The following day samples were fragmented for 1hour at 37 degrees, precipitated at 4 degrees for 30 minutes, and pelleted at 4 degrees for 20 minutes at 3000xg. Samples were dried in a hood at room temperature for 1 hour and re-suspended in the recommended volume of RA1 following Illumina’s Infinium HD assay at 48 degrees Celsius for 1 hour. Samples were then denatured at 95 degrees Celsius for 20 minutes. Samples had a 10-minute cool down and then directly hybridized to Illumina Methylation Epic Array Cat. No. WG-317-1001. Sample placement in each chip was randomized. Hybridization was for 18 hours at 48 degrees Celsius using a stabilized hybridization oven. The following day arrays were washed and stained automatically using the Tecan Freedom Evo technology. Arrays were then be dipped in the UV protectant (Illumina’s XC3) for ten minutes and any excess removed. Arrays were dried for 1 hour in a vacuum desiccator. Arrays were scanned using Illumina’s iScan array scanner and raw files generated. Raw data files were imported into Illumina’s GenomeStudio software and a project created as well as QC parameters checked to ensure project went as expected. Scanned output files were analyzed and call rates calculated using Illumina’s GenomeStudio. The quality of the data was evaluated by both sample dependent and sample independent controls. Specifically, efficiency of target removal, non-specific binding, and appearance of cross-contamination was examined. All raw Data and the GenomeStudio project were uploaded to the password protected Keck Microarray Database.

#### Data Analysis

DNAmAge was calculated using the online Horvath clock available at https://dnamage.genetics.ucla.edu/. Analysis of epigenetic age was performed, blinded, on the final 18 participants in the treatment group and 20 participants in the control group. P values were computed as an unpaired 2-tailed t-test between the experimental group and control group, using the individual score differences (after treatment minus before) as a random variable.

#### Data Sharing

The data that support the findings will be available in Gene Expression Omnibus at https://www.ncbi.nlm.nih.gov/geo/, submission number GSE 149747, from 04-14-23 following an embargo from the date of publication to allow for commercialization of research findings.

## 5 ACKNOWLEDGMENTS

We would like to acknowledge and thank the following for their assistance in this research study: The team at the National University of Natural Medicine Helfgott Research Institute for conducting the clinical trial. Kari Thostensen and MBody360 for their collaboration and the use of their coaching platform. Marquelle Brown, Janine Henkel, Alison Parkerson and Brianne Pugh for their contributions to diet-compatible recipe development. Yale University for measuring CpG site methylation according to the Illumina Infinium array. Dr. Moshe Szyf’s team at the Department of Pharmacology and Therapeutics at McGill University for their input on data analysis. Josh Mitteldorf for his extensive assistance with data analysis. Steve Horvath PhD ScD, for input on the calculation of epigenetic age.

This study was generously supported through an unrestricted grant from Metagenics, Inc.

## 7 AUTHOR CONTRIBUTIONS

KF, RB and MS designed the study. RH and KF designed the study diet. RB and ES conducted the clinical trial at Helfgott Research Institute. JH, SL, JH, DG and MT coached the participants through the intervention. DH performed the data analysis and reporting with MS, DC and DH in an advisory capacity. KF, RB and RH developed the manuscript. All authors approved the manuscript.

## REFERENCES

Barja, G. (2019). Towards a unified mechanistic theory of aging. Experimental gerontology, 124, 110627.

Boyd-Kirkup, J. D., Green, C. D., Wu, G., Wang, D., & Han, J.-D. J. (2013). Epigenomics and the regulation of aging. Epigenomics, 5(2), 205–227.

Carroll, J. E., Irwin, M. R., Levine, M., Seeman, T. E., Absher, D., Assimes, T., & Horvath, S. (2017). Epigenetic aging and immune senescence in women with insomnia symptoms: findings from the Women’s Health Initiative Study. Biological psychiatry, 81(2), 136–144.

Carskadon, M. A., Chappell, K. R., Barker, D. H., Hart, A. C., Dwyer, K., Gredvig-Ardito, C., … McGeary, J. E. (2019). A pilot prospective study of sleep patterns and DNA methylation-characterized epigenetic aging in young adults. BMC research notes, 12(1), 1–5.

Chen, B. H., Marioni, R. E., Colicino, E., Peters, M. J., Ward-Caviness, C. K., Tsai, P.-C., … Horvath, S. (2016). DNA methylation-based measures of biological age: meta-analysis predicting time to death. Aging (Albany NY), 8(9), 1844–1859. doi:10.18632/aging.101020

de Grey, A. D. (2005). The unfortunate influence of the weather on the rate of ageing: why human caloric restriction or its emulation may only extend life expectancy by 2-3 years. Gerontology, 51(2), 73–82. doi:10.1159/000082192

e Silva, A. d. S., & da Mota, M. P. G. (2014). Effects of physical activity and training programs on plasma homocysteine levels: a systematic review. Amino acids, 46(8), 1795–1804.

Ebbing, M., Bonaa, K. H., Nygard, O., Arnesen, E., Ueland, P. M., Nordrehaug, J. E., … Vollset, S. E. (2009). Cancer incidence and mortality after treatment with folic acid and vitamin B12. JAMA, 302(19), 2119–2126. doi:10.1001/jama.2009.1622

Fahy, G. M., Brooke, R. T., Watson, J. P., Good, Z., Vasanawala, S. S., Maecker, H., … Horvath, S. (2019). Reversal of epigenetic aging and immunosenescent trends in humans. Aging cell.

Field, A. E., Robertson, N. A., Wang, T., Havas, A., Ideker, T., & Adams, P. D. (2018). DNA methylation clocks in aging: categories, causes, and consequences. Molecular cell, 71(6), 882–895.

Figueiredo, J. C., Grau, M. V., Haile, R. W., Sandler, R. S., Summers, R. W., Bresalier, R. S., … Baron, J. A. (2009). Folic acid and risk of prostate cancer: results from a randomized clinical trial. J Natl Cancer Inst, 101(6), 432–435. doi:10.1093/jnci/djp019

Goldman, D. (2017). THE ECONOMIC RETURNS TO DELAYED AGING: PROMISE AND PITFALLS. Innovation in Aging, 1(Suppl 1), 1082.

Hariri, M., Salehi, R., Feizi, A., Mirlohi, M., Ghiasvand, R., & Habibi, N. (2015). A randomized, double-blind, placebo-controlled, clinical trial on probiotic soy milk and soy milk: effects on epigenetics and oxidative stress in patients with type II diabetes. Genes & nutrition, 10(6), 52.

Hayano, M., Yang, J.-H., Bonkowski, M. S., Amorim, J. A., Ross, J. M., Coppotelli, G., … Yang, X. (2019). DNA Break-Induced Epigenetic Drift as a Cause of Mammalian Aging.

Hore, T. A. (2017). Modulating epigenetic memory through vitamins and TET: implications for regenerative medicine and cancer treatment. Epigenomics, 9(6), 863–871.

Horvath, S. (2013). DNA methylation age of human tissues and cell types. Genome biology, 14(10), R115.

Horvath, S., Oshima, J., Martin, G. M., Lu, A. T., Quach, A., Cohen, H., … Kabacik, S. (2018). Epigenetic clock for skin and blood cells applied to Hutchinson Gilford Progeria Syndrome and ex vivo studies. Aging (Albany NY), 10(7), 1758.

Horvath, S., & Raj, K. (2018). DNA methylation-based biomarkers and the epigenetic clock theory of ageing. Nature Reviews Genetics, 1.

Jin, K., Simpkins, J. W., Ji, X., Leis, M., & Stambler, I. (2015). The critical need to promote research of aging and aging-related diseases to improve health and longevity of the elderly population. Aging and disease, 6(1), 1.

Johnson, A. A., Akman, K., Calimport, S. R., Wuttke, D., Stolzing, A., & de Magalhães, J. P. (2012). The Role of DNA Methylation in Aging, Rejuvenation, and Age-Related Disease. Rejuvenation Research, 15(5), 483–494.

Kane, A. E., & Sinclair, D. A. (2019). Epigenetic changes during aging and their reprogramming potential. Critical reviews in biochemistry and molecular biology, 54(1), 61–83.

Langie, S. A. S., Moisse, M., Declerck, K., Koppen, G., Godderis, L., Vanden Berghe, W., … De Boever, P. (2017). Salivary DNA Methylation Profiling: Aspects to Consider for Biomarker Identification. Basic Clin Pharmacol Toxicol, 121 Suppl 3, 93-101. doi:10.1111/bcpt.12721

Levine, M. E., Lu, A. T., Quach, A., Chen, B. H., Assimes, T. L., Bandinelli, S., … Li, Y. (2018). An epigenetic biomarker of aging for lifespan and healthspan. Aging (Albany NY), 10(4), 573.

Li, E., & Zhang, Y. (2014). DNA methylation in mammals. Cold Spring Harbor perspectives in biology, 6(5), a019133.

Lorusso, J. S., Sviderskiy, O. A., & Labunskyy, V. M. (2018). Emerging Omics Approaches in Aging Research. Antioxid Redox Signal, 29(10), 985–1002. doi:10.1089/ars.2017.7163

Mitteldorf, J. (2013). How does the body know how old it is? Introducing the epigenetic clock hypothesis. Biochemistry (Moscow), 78(9), 1048–1053.

Moore, S. R., McEwen, L. M., Quirt, J., Morin, A., Mah, S. M., Barr, R. G., … Kobor, M. S. (2017). Epigenetic correlates of neonatal contact in humans. Development and psychopathology, 29(5), 1517–1538.

Nilsson, E. K., Boström, A. E., Mwinyi, J., & Schiöth, H. B. (2016). Epigenomics of total acute sleep deprivation in relation to genome-wide DNA methylation profiles and RNA expression. Omics: a journal of integrative biology, 20(6), 334–342.

Obeid, R., Hubner, U., Bodis, M., Graeber, S., & Geisel, J. (2018). Effect of adding B-vitamins to vitamin D and calcium supplementation on CpG methylation of epigenetic aging markers. Nutr Metab Cardiovasc Dis, 28(4), 411–417. doi:10.1016/j.numecd.2017.12.006

Oliai Araghi, S., Kiefte-de Jong, J. C., van Dijk, S. C., Swart, K. M. A., van Laarhoven, H. W., van Schoor, N. M., … van der Velde, N. (2019). Folic Acid and Vitamin B12 Supplementation and the Risk of Cancer: Long-term Follow-up of the B Vitamins for the Prevention of Osteoporotic Fractures (B-PROOF) Trial. Cancer Epidemiol Biomarkers Prev, 28(2), 275–282. doi:10.1158/1055-9965.EPI-17-1198

Pal, S., & Tyler, J. K. (2016). Epigenetics and aging. Science advances, 2(7), e1600584.

Panel, C. C., Watson, N. F., Badr, M. S., Belenky, G., Bliwise, D. L., Buxton, O. M., … Grandner, M. A. (2015). Joint consensus statement of the American Academy of Sleep Medicine and Sleep Research Society on the recommended amount of sleep for a healthy adult: methodology and discussion. Sleep, 38(8), 1161–1183.

Pauwels, S., Ghosh, M., Duca, R. C., Bekaert, B., Freson, K., Huybrechts, I., … Godderis, L. (2017). Maternal intake of methyl-group donors affects DNA methylation of metabolic genes in infants. Clin Epigenetics, 9. doi:10.1186/s13148-017-0321-y

Pavanello, S., Campisi, M., Tona, F., Dal Lin, C., & Iliceto, S. (2019). Exploring Epigenetic Age in Response to Intensive Relaxing Training: A Pilot Study to Slow Down Biological Age. International journal of environmental research and public health, 16(17), 3074.

Quach, A., Levine, M. E., Tanaka, T., Lu, A. T., Chen, B. H., Ferrucci, L., … Beasley, J. M. (2017). Epigenetic clock analysis of diet, exercise, education, and lifestyle factors. Aging (Albany NY), 9(2), 419.

Rando, T. A., & Chang, H. Y. (2012). Aging, rejuvenation, and epigenetic reprogramming: resetting the aging clock. Cell, 148(1), 46–57.

Ren, H., Collins, V., Clarke, S. J., Han, J.-S., Lam, P., Clay, F., … Andy Choo, K. (2012). Epigenetic changes in response to tai chi practice: a pilot investigation of DNA methylation marks. Evidence-based Complementary and Alternative Medicine, 2012.

Sae-Lee, C., Corsi, S., Barrow, T. M., Kuhnle, G. G. C., Bollati, V., Mathers, J. C., & Byun. (2018).Dietary Intervention Modifies DNA Methylation Age Assessed by the Epigenetic Clock. Molecular nutrition & food research, 62(23). doi:10.1002/mnfr.201800092

Sen, P., Shah, P. P., Nativio, R., & Berger, S. L. (2016). Epigenetic mechanisms of longevity and aging. Cell, 166(4), 822–839.

Spólnicka, M., Pośpiech, E., Adamczyk, J. G., Freire-Aradas, A., Pepłońska, B., Zbieć-Piekarska, R., … Phillips, C. (2018). Modified aging of elite athletes revealed by analysis of epigenetic age markers. Aging (Albany NY), 10(2), 241.

Sybesma, W., Starrenburg, M., Tijsseling, L., Hoefnagel, M. H., & Hugenholtz, J. (2003). Effects of cultivation conditions on folate production by lactic acid bacteria. Appl. Environ. Microbiol., 69(8), 4542–4548.

Tu, H., Dinney, C. P., Ye, Y., Grossman, H. B., Lerner, S. P., & Wu. (2018). Is Folic Acid Safe for Non-Muscle-Invasive Bladder Cancer Patients? An Evidence-Based Cohort Study. The American journal of clinical nutrition, 107(2). doi:10.1093/ajcn/nqx019

van Wijngaarden, J. P., Swart, K. M., Enneman, A. W., Dhonukshe-Rutten, R. A., van Dijk, S. C., Ham, A. C., … de Groot, L. C. (2014). Effect of daily vitamin B-12 and folic acid supplementation on fracture incidence in elderly individuals with an elevated plasma homocysteine concentration: B-PROOF, a randomized controlled trial. Am J Clin Nutr, 100(6), 1578–1586. doi:10.3945/ajcn.114.090043

Waterland, R. A., & Jirtle, R. L. (2003). Transposable elements: targets for early nutritional effects on epigenetic gene regulation. Molecular and cellular biology, 23(15), 5293–5300.

West, M. D., Sternberg, H., Labat, I., Janus, J., Chapman, K. B., Malik, N. N., … Larocca, D. (2019). Toward a unified theory of aging and regeneration. Regenerative medicine, 14(9), 867–886.

White, A. J., Sandler, D. P., Bolick, S. C., Xu, Z., Taylor, J. A., & DeRoo, L. A. (2013). Recreational and household physical activity at different time points and DNA global methylation. European journal of cancer, 49(9), 2199–2206.

Wolf, E. J., Logue, M. W., Hayes, J. P., Sadeh, N., Schichman, S. A., Stone, A., … Miller, M. W. (2016). Accelerated DNA methylation age: associations with PTSD and neural integrity. Psychoneuroendocrinology, 63, 155–162.

Zannas, A. S., Arloth, J., Carrillo-Roa, T., Iurato, S., Röh, S., Ressler, K. J., … Heim, C. (2015). Lifetime stress accelerates epigenetic aging in an urban, African American cohort: relevance of glucocorticoid signaling. Genome biology, 16(1), 266.

Zhong, J., Karlsson, O., Wang, G., Li, J., Guo, Y., Lin, X., … Baccarelli, A. A. (2017). B vitamins attenuate the epigenetic effects of ambient fine particles in a pilot human intervention trial. Proc Natl Acad Sci U S A, 114(13), 3503–3508. doi:10.1073/pnas.1618545114

